# Prevalence of SARS-CoV-2 IgG antibodies in a population from Veracruz (Southeastern Mexico)

**DOI:** 10.1101/2020.10.19.20215558

**Authors:** Jose María Remes-Troche, Antonio Ramos-De-la-Medina, Marisol Manríquez-Reyes, Laura Martínez-Pérez Maldonado, María Antonieta Solis-Gonzalez, Karina Guadalupe Hernández Flores, Héctor Vivanco Cid

**Affiliations:** Medical Biological Research Institute, Universidad Veracruzana, Veracruz, México; Hospital Español de Veracruz, Veracruz, México

**Author notes:** Corresponding author: Dr José María Remes-Troche. Instituto de Investigaciones Médico-Biológicas, Universidad Veracruzana, Veracruz. Av. Iturbide s/n Entre Carmen Serdán y 20 de Nov. Col. Centro, C.P. 91700 Veracruz, Ver. México. Specific author contributions: Study concept and design – Remes-Troche JM, Ramos-De-la-Medina A, Vivanco-Cid H. Data Acquisition – Laura Martínez-Pérez Maldonado, Antonieta Solís-González, Karina Guadalupe Hernández Flores. Paper preparation and statistical analysis – Remes-Troche JM, Ramos-De-la Medina A, Marisol Manríquez-Reyes, Héctor Vivanco-Cid. Critical revisions – Remes-Troche JM, Ramos-De-la Medina A, Héctor Vivanco-Cid. Administrative support and overall study supervision –Remes-Troche JM. Funding: None.

**Keywords:** SARS-CoV-2, COVID-19, seroprevalence, Mexico, antibodies

## Abstract

**Introduction/Aim:** Recent studies have shown that seroprevalence is quite variable depending on the country, the population and the time of the pandemic in which the serological tests are performed. Here, we investigated the prevalence of IgG antibodies against SARS-CoV-2 in a population living in Veracruz City, México.

**Methods:** From of June 1 to July 31, 2020, the consecutive adult patients (age ≥18 years) that attended 2 ambulatory diagnostic private practice centers for testing were included. Samples were run on the Abbott Architect instrument using the commercial Abbott SARS-CoV-2 IgG assay. The main outcome was seroprevalence. Demographics, previous infection to SARS-CoV-2 (according to a previous positive polymerase-chain reaction nasopharyngeal swab), self-suspicious of virus of infection (according to have in the previous 4 weeks either fever, headache, respiratory symptoms but not a confirmatory PCR) or no having symptoms were also evaluated.

**Results:** A total of 2174 subjects were tested, included 53.6% women (mean age 41.8±15.17 years, range 18-98 years). One thousand and forty-one (52.5%) subjects were asymptomatic, 722 (33.2%) had suspicious of infection and 311 (14.3%) had previous infection. Overall, 642 of 2174 (29.5% [95% CI 27.59%-31.47%]) of our population were seropositive. Seropositivity among groups was 21.3% in asymptomatic, 23.4% in self-suspicious patients and 73.9% in previous infection patients.

**Conclusions:** We found one of the highest seroprevalences reported for SARS-CoV-2 worldwide in asymptomatic subjects (21.3%) as well in subjects with self-suspicious of COVID-19 (23.4%). The number of infected subjects in our population is not encouraging and it should be interpreted with caution.

## Introduction

Currently, Latin America is the epicenter of the COVID 19 pandemic, and Mexico is one of the most affected countries being the seventh country (730,317) with the most cases and the fourth (76,430) in mortality as of September 28th, 2020. Population-based seroprevalence measuring anti-severe acute respiratory syndrome coronavirus 2 (anti-SARS-CoV-2) antibodies provide one method for estimating infection rates and monitoring the progression of the epidemic.^1^ Recent studies have shown that seroprevalence is quite variable depending on the country, the population and the time of the pandemic in which the serological tests are performed.^2,3,4,5,6^

Here, we investigated the prevalence of IgG antibodies against SARS-CoV-2and associations with demographics, previous SARS-CoV-2 infection or suspicion of virus exposure, as well in asymptomatic subjects.

## Methods

From of June 1 to July 31, 2020, the consecutive adult patients (age ≥18 years) that attended 2 ambulatory diagnostic private practice centers for testing were included. Samples were run on the Abbott Architect instrument using the commercial Abbott SARS-CoV-2 IgG assay. The main outcome was seroprevalence. Seroprevalence with 95% confidence interval was calculated by the exact binomial technique. Demographics, previous infection to SARS-CoV-2 (according to a previous positive polymerase-chain reaction nasopharyngeal swab), self-suspicious of virus of infection (according to have in the previous 4 weeks either fever, headache, respiratory symptoms but not a confirmatory PCR) or no having symptoms were also evaluated. Associations among seroprevalence and these variables was assessed using chi-square analysis and logistic regression. IBM® SPSS Statistics® version 22 was used for analyses. *P* < .05 (2-sided) defined statistical significance. The Hospital Español institutional review board approved this research.

## Results

A total of 2174 subjects were tested, included 53.6% women (mean age 41.8±15.17 years, range 18-98 years). One thousand and forty-one (52.5%) subjects were asymptomatic, 722 (33.2%) had suspicious of infection and 311 (14.3%) had previous infection. Overall, 642 of 2174 (29.5% [95% CI 27.59%-31.47%]) of our population were seropositive. Seropositivity among groups was 21.3% (n=243/1141 [95% CI 18.78%-23.71%]) in asymptomatic, 23.4% (n=230/311 [95% CI 20.24%-26.56%])in self-suspicious patients and 73.9% (n=230/311 [95% CI 68.91%-78.99%])in previous infection patients (p <0.0001). In the bivariate analysis male sex (p=0.014, RR 1.17 [95% CI 1.03-1.33]) and age (p=0.0017) were associated to seroprevalence (Table). In the multivariate analysis age (p=0.016) was the only variable associated to seroprevalence

## Discussion

The number of infected subjects in our Mexican population is not encouraging and it should be interpreted with caution. Although some might think that we can be closer to the “so-called” herd immunity, on the other hand it could also make us think that in the case of reinfection our situation would not be so promising, since it be also higher. We fully understand that the true seroprevalence to SARS-CoV-2 is difficult to estimate at this point in the pandemic and with current essays.

Our study has obvious limitations. Selection bias is likely. The estimated prevalence may be biased due to symptomatic individuals and their family members may have been more likely to participate. Prevalence estimates could change with new information on the accuracy of test kits used. Also, the study was limited to our region of our county. Serologic testing in other locations is warranted to track the progress of the epidemic in the whole country.

**Table 1.** Demographic Information and Seroprevalence for 2,159Subjects Tested for COVID-19 IgG Antibodies in the Veracruz city.

|  | Seropositive to IgG<br>SARS-CoV-2 | Seronegative to IgG<br>SARS-nCoV-2 | p value | OR (CI 95%) |
| --- | --- | --- | --- | --- |
|  | n= 642 | n=1532 |  |  |
| Sex (n,%) |  |  |  |  |
| - Female | 318 (49.5%) | 847 (55.3%) | 0.014 | 1.26 (1.047-1.515) |
| - Male | 324 (50.5%) | 685 (44.7%) |  |  |
| Age (mean ± SD) | 41.17 ± 15 | 48.2 ± 14 | 0.0001 | ----- |
| Age group (n,%) |  |  |  |  |
| < 29 | 115 (17.9%) | 388 (25.3%) | 0.0017 | 1.0 |
| 30-39 | 176 (27.4%) | 378 (24.7%) |  | 1.57 (1.19-2.06) |
| 40-49 | 131 (20.4%) | 339 (22.1%) |  | 1.30 (0.97-1.74) |
| 50-59 | 116 (18.1%) | 228 (14.9%) |  | 1.71 (1.26-2.32) |
| 60-69 | 77 (12%) | 128 (8.4%) |  | 2.02 (1.42-2.88) |
| > 70 | 27 (4.2%) | 71 (4.6%) |  | 1.28 (0.78-2.09) |

## Data Availability

If requested

## References

1.- Eckerle I, Meyer B. SARS-CoV-2 seroprevalence in COVID-19 hotspots [published online ahead of print, 2020 Jul 3]. Lancet. 2020;396(10250):514–515. doi:10.1016/S0140-6736(20)31482-3

2.- Pollán M, Pérez-Gómez B, Pastor-Barriuso R. Prevalence of SARS-CoV-2 in Spain (ENE-COVID): a nationwide, population-based seroepidemiological study. Lancet. 2020 doi: 10.1016/S0140-6736(20)31483-5. published online July 6

3.- Stringhini S, Wisniak A, Piumatti G. Seroprevalence of anti-SARS-CoV-2 IgG antibodies in Geneva, Switzerland (SEROCoV-POP): a population-based study. Lancet. 2020 doi: 10.1016/S0140-6736(20)31304-0. published online June 11

4.- Sood N, Simon P, Ebner P, et al. Seroprevalence of SARS-CoV-2-Specific Antibodies Among Adults in Los Angeles County, California, on April 10-11, 2020 [published online ahead of print, 2020 May 18]. JAMA. 2020;323(23):2425–2427. doi:10.1001/jama.2020.8279

5.- Fiore JR, Centra M, De Carlo A, et al. Results from a survey in healthy blood donors in South Eastern Italy indicate that we are far away from herd immunity to SARS-CoV-2 [published online ahead of print, 2020 Aug 13]. J Med Virol. 2020;10.1002/jmv.26425. doi:10.1002/jmv.26425

6.- Moscola J, Sembajwe G, Jarrett M, et al. Prevalence of SARS-CoV-2 Antibodies in Health Care Personnel in the New York City Area [published online ahead of print, 2020 Aug 6]. JAMA. 2020;e2014765. doi:10.1001/jama.2020.14765

